# Self-reported Xylazine Experiences: A Mixed Methods Study of Reddit Subscribers

**DOI:** 10.1101/2023.03.13.23287215

**Authors:** Anthony Spadaro, Karen O’Connor, Sahithi Lakamana, Abeed Sarker, Rachel Wightman, Jennifer S Love, Jeanmarie Perrone

**Author notes:** Corresponding author: Anthony Spadaro, Hospital of the University of Pennsylvania, 3400 Spruce Street, Philadelphia, PA, 19104.

## Abstract

**Objectives:** Xylazine is an alpha-2 agonist increasingly prevalent in the illicit drug supply. Our objectives were to curate information about xylazine through social media from People Who Use Drugs (PWUDs). Specifically, we sought to answer the following: 1) what are the demographics of Reddit subscribers reporting exposure to xylazine? 2) is xylazine a desired additive? and 3) what adverse effects of xylazine are PWUDs experiencing?

**Methods:** Natural Language Processing (NLP) was used to identify mentions of “xylazine” from posts by Reddit subscribers who also posted on drug-related subreddits. Posts were qualitatively evaluated for xylazine-related themes. A survey was developed to gather additional information about the Reddit subscribers. This survey was posted on subreddits that were identified by NLP to contain xylazine-related discussions from March 2022 to October 2022.

**Results:** 76 posts mentioning xylazine were extracted via NLP from 765,616 posts by 16,131 Reddit subscribers (January 2018 to August 2021). People on Reddit described xylazine as an unwanted adulterant in their opioid supply. 61 participants completed the survey. Of those that disclosed their location, 25/50 (50%) participants reported locations in the Northeastern United States. The most common eoute of xylazine use was intranasal use (57%). 31/59 (53%) reported experiencing xylazine withdrawal. Frequent adverse events reported were prolonged sedation (81%) and increased skin wounds (43%).

**Conclusions:** Among respondents on these Reddit forums, xylazine appears to be an unwanted adulterant. PWUDs may be experiencing adverse effects such as prolonged sedation and xylazine withdrawal. This appeared to be more common in the Northeast.

## Introduction

Xylazine is an α_2_-agonist used in veterinary medicine as a sedative-hypnotic, muscle relaxant, and anti-emetic.^1^ It is not approved for human use. Xylazine was investigated for potential use in humans, but rejected due to severe central nervous system and blood pressure adverse effects^2–4^. The chemical structure of xylazine resembles phenothiazines, tricyclic antidepressants, and clonidine.^1,5^ In addition to sedative effects caused by central A2 agonism there is evidence that xylazine acts at multiple receptors contributing to its clinical effects..^1,4,5^ Veterinary studies have evaluated its use as a sedative in a number of different species.^6,7^ Until recently, descriptions of human exposures to xylazine in the medical literature were limited to case reports of xylazine intentional exposures.^4,8–10^ There are sporadic reports of xylazine being present in the illicit drug supply in Philadelphia and Puerto Rico prior to 2019.^11,12^

Emerging evidence from forensic, confiscated, and donated drug samples demonstrate that xylazine is increasingly found in the opioid drug supply in the Northeast and rapidly spreading to the rest of the country.^2,13–15^ In 2019, the CDC reported that of 45,676 overdose deaths in the State Unintentional Drug Overdose Reporting System (SUDORS) there were 826 (1.8%) that were xylazine-positive, and that 99.1% of all xylazine-involved deaths also involved fentanyl or fentanyl analogues.^16^ More recently in Philadelphia, xylazine is being found in up to 75-90% of drug samples.^15^ Despite its increasing prevalence, few studies have described the limited qualitative experience of concurrent xylazine use with opioids among people who use drugs (PWUDS).^14^

Reddit is one of the most popular and fastest-growing social networks currently, with over 430 million monthly active subscribers.^17^ It allows subscribers to remain completely anonymous if they choose, and, due to this anonymity, Reddit has become a hub for the open discussion of sensitive or stigmatized topics including drug use.^18,19^ Given rising concerns about xylazine we sought to identify a cohort of people who self report exposure to xylazine in the drug supply by using public social media data from Reddit to conduct this study.^19,20^ Our objectives were to better understand xylazine use by utilizing social media to survey PWUDs about their experience with xylazine. Specifically, we sought to answer the following: 1) what are the demographics of Reddit subscribers self-reporting exposure to xylazine? 2) is xylazine a desired additive? and 3) what adverse effects of xylazine are PWUDs experiencing?

## Methods

This is a mixed methods study of PWUDs who use Reddit. Study methods include natural language processing (NLP) to identify posts on Reddit related to xylazine use for qualitative analysis as well as posting a survey to Reddit to elicit the experiences of PWUD for quantitative analysis.

### Qualitative analysis of existing posts

Several drug-related subreddits on Reddit were selected to search for xylazine-related posts. Initial subreddits were selected based on study group consensus of subreddit relevance and prior experience using Reddit to evaluate patterns in drug use.^19^ From the chosen subreddits, we collected all publicly available retrievable posts via the PRAW Python application programming interface (API) for Reddit.^21^ We searched for mentions of ‘xylazine’ and ‘Tranq’, as well as lexical variants, from the subreddits using NLP to identify references to xylazine.^22^ Two authors (K.O. and A.S.) reviewed a small sample of posts to identify relevant categories into which the posts could be coded. We used a thematic content analysis approach to classify posts into different codes.^23,24^ A consensus approach among the authors was used to assign codes to the posts. The coding process was a multi-label classification and each post could be assigned one or many codes.^23^

### Survey Design

An anonymous English-language survey was developed to include questions about demographics, patterns of xylazine use, and experiences from using xylazine. The survey was created on the survey hosting platform Qualtrics (https://www.qualtrics.com/) which allowed for web-based distribution of survey via a link. All respondents gave digital informed consent prior to beginning the survey. Survey questions did not include any personally identifiable information or contact information in order to preserve the respondents’ anonymity. The survey was 18 questions and there was no financial incentive (Supplement S1). The survey was posted on seven drug-related subreddits from March 2022 to August 2022 by a member of our research team (A.S.), who identified himself and the goal of the study on all of the posts that included the link to the survey, and obtained consent from the moderators (Supplement S2). The subreddits where the survey was posted were identified through the qualitative analysis as having the highest number of posts about xylazine. Additional drug related subreddits with large numbers of subscribers were also identified as potential locations to post the survey. From this set, only subreddits where the moderators allowed the posting of the survey were included. The survey link was posted after the qualitative data had been gathered so as to not influence the content of posts being analyzed in the qualitative analysis. The anonymous survey results were exported to Microsoft Excel for analysis.

### Ethics Approval

This study was approved by XXX Institutional Review Board (IRB) #: STUDY00002458.

## Results

### NLP Results

15 subreddits were found to contain at least one possible reference to the substance (*r/fentanyl, r/heroin, r/researchchemicals, r/OpiateChurch, r/ObscureDrugs, r/HeroinHeroines, r/ayahuasca, r/Methadone, r/OurOverUsedVeins, r/pillreports, r/popping, r/morbidquestions, r/noids, r/Drugs, r/DrugsOver30*). Possible misspellings of xylazine were also generated via a validated NLP tool prior to conducting the search.^22^ Seventy six posts were identified through NLP among these subreddits. Fourteen posts were excluded for 1) not mentioning xylazine in the post or 2) discussing a veterinary use of xylazine. Major themes that were identified included discussion of other drugs used with xylazine, method of using xylazine, and geography of xylazine use. Additionally, posts that discussed dependence or addiction to xylazine and other drugs were identified to further understand patterns of xylazine use as well as to explore the impact xylazine might have on medications for opioid use disorder (MOUDs). Table 1 includes major themes with example quotes and counts of times major themes appeared.

**Table 1:** Major themes from Reddit posts about Xylazine

| Theme | Definition | Example Quotes | Theme Frequency<br>(% of posts theme<br>appeared in) |
| --- | --- | --- | --- |
| Other Drugs used<br>with Xylazine | Posts that<br>discussed xylazine<br>use with other<br>specific drugs | “Around me it's usually<br>fetty/etonitazene analogs<br>mixed with Xylazine which is<br>an animal tranq similar<br>chemically to clonidine, and<br>sometimes ketamine is<br>thrown in too alongside<br>synthetic cannabinoids” | 53% |
| Sentiment about<br>Xylazine | Posts that<br>described a positive<br>or negative<br>experience about<br>using xylazine | “I fucking LOVED xylazine... I<br>loved it. I was so upset if my<br>fetty wasn't cut with it.” | 24% |
| Method of<br>Xylazine use | Posts that<br>described how<br>xylazine was used | “Always break the tiniest<br>crumb off to test new<br>batches. Smoking helps<br>cause it hits pretty fast so<br>you can judge” | 6% |
| Geographic Use | Posts that<br>described a<br>geographic area | “It's called xylazine... philly's<br>got that "tranq dope".. puts<br>you out but you get addicted<br>to that along with the fent!” | 37% |
| Dependence on<br>xylazine and other<br>substances | Posts that discuss<br>dependence,<br>addiction, or<br>withdrawal to<br>xylazine and other<br>substances | “12 hours later from my last<br>shot of this<br>fent/trank/ketamine/xylazine<br>shit i started going thrpigh<br>the worst withdraws ever.<br>drank another 115<br>methadone and within 15<br>minutes thru it up<br>everywhere. withdraw is<br>ridiculous and methadone<br>doesnt fucking cut it” | 11% |
| Use of MOUDs | Posts that discuss<br>buprenorphine,<br>methadone, | “I'm on 80 mgs of<br>methadone .. we started<br>getting weekly take home | 2% |
|  | naltrexone or other MOUDS | bottles from this covid shit do I would get a few bags here or there. Thinking ahh I got the methadone to fall back on an I'll be fine.. well come to find out the shit is heavily cut with xylazine” |  |

### Survey Results

The survey link was posted 17 times in total on the subreddits *r/*opiates, *r/*fentanyl, *r/*DrugNerds, *r/*ObscureDrugs, *r/*researchchemicals, *r/heroin, r/suboxone*. There were a total of 61 participants who completed the survey. Survey participant demographics are outlined in Table 2. The median age range reported by participants was 30-39 years old. The majority were male (67%), white non-Hispanic (79%), and from the Northeast United States (50%). Of the 25 responses that were included from a state in the Northeast, 12 responses were specifically from Pennsylvania.

**Table 2.** Demographic Results of survey participants

|  |  |
| --- | --- |
| **Age Range (N=51)** |  |
| **Under 20** | 8% |
| **20-29** | 33% |
| **30-39** | 47% |
| **40-49** | 12% |
| **Over 50** | 0% |
| **Gender (N=51)** |  |
| Male | 67% |
| Female | 29% |
| Non-Binary/Other | 4% |
| **Race (N=50)** |  |
| White, Non-Hispanic | 79% |
| White, Hispanic | 4% |
| African American | 4% |
| Asian | 5% |
| American Indian | 4% |
| Native Hawaiian/Pacific Islander | 4% |
| **Geography (N=50)** |  |
| Northeast | 50% |
| Southeast | 14% |
| Midwest | 14% |
| Southwest | 0% |
| West | 10% |
| Other Country* | 12% |

\*Included 2 responses from the United Kingdom, 2 responses from Canada, 1 response from Sweden, and 1 response from Ecuador.

Participants were asked about drug use history including frequency of use, routes of exposure, and concurrent drug use with xylazine (Table 3). A majority of participants (74%) report that they do not intentionally seek out xylazine. Most of the participants used multiple other drugs, with fentanyl specifically being the most common opioid (80%). Many participants used multiple different methods of xylazine with intranasal (57%) and injection (43%) being the most common.

**Table 3.** Patterns of Xylazine Use

|  |  |
| --- | --- |
| **Intentionally seek out to buy drugs that contain xylazine (N=61)** |  |
| Yes | 26% |
| No | 74% |
| **Drug use on a typical day (N=60)** |  |
| Any Opioids | 93% |
| Fentanyl | 80% |
| Cannabis | 45% |
| Stimulants | 41% |
| Benzodiazepines | 21% |
| Hallucinogens | 10% |
| **Method of xylazine exposure (N=61)** |  |
| Inhalational (Smoking) | 20% |
| Intranasal (Snorting) | 57% |
| Injection | 43% |
| Oral | 3% |
| **Frequency of xylazine use (N=61)** |  |
| Daily | 39% |
| 1-6 times per week | 19% |
| 1-4 times per month | 15% |
| A few times per year/Rarely | 27% |

Participants reported adverse effects from using xylazine (Table 4). A majority of participants (53%) reported having withdrawal from using xylazine. Common symptoms of withdrawal included anxiety (91%), depressed mood (74%), and body aches (63%). A majority (57%) of patients reported that their xylazine use has made withdrawing from other substances worse.

**Table 4:** Reported Adverse Effects of Xylazine use

|  |  |
| --- | --- |
| **Adverse Effects from xylazine use (N=48)** |  |
| Increased overdoses/passing out | 81% |
| Skin Wounds or Infections | 43% |
| Increased Emergency Room visits | 17% |
| **Have you experienced withdrawal from xylazine? (N=59)** |  |
| Yes | 53% |
| **What xylazine withdrawal symptoms have you experienced (N=35)** |  |
| Body Aches | 63% |
| Cravings | 49% |
| Anxiety | 91% |
| Depressed Mood | 74% |
| **How has withdrawing from other drugs changed since you started using xylazine? (N=51)** |  |
| Worse | 57% |
| The Same | 39% |
| Better | 4% |

The participants emphasized many of these adverse effects of xylazine in a free response prompt to share any specifics about xylazine. Some participants emphasized the severity of the wounds they experienced as:

“*bilateral purulent skin ulcers in extremities, skin infections, tissue death*.”

“*The wounds last months my primary has had me on 3 rounds of antibiotics with no luck*”.

Other participants commented that xylazine made their opioid withdrawal worse:

“*Suboxone an methadone don’t help if there is tranq in your dope*”.

Several participants comment on autonomic effects that weren’t asked about in the survey such as labile blood pressure:

“*Very high blood pressure, can feel heart beat in your face its so bad. Chest pains. Stretching feeling*.”

“*Coming off tranq makes your blood pressure spike with dizziness an lips go numb*”.

## Discussion

This novel study adds to the literature about xylazine, an emerging psychoactive substance that is increasingly being detected in the illicit drug supply. This study found that most of the participants did not seek out xylazine, participants were primarily from the northeastern United States, and many participants reported increased wounds and worsened withdrawal symptoms.

First, we found that most participants were not intentionally seeking out xylazine in the drugs they use daily. Other studies of xylazine have found it to be an undesired adulterant in the drug supply.^14,25^ One notable finding from this study was that there was a group of participants that reported that they intentionally sought out xylazine. This behavior may be motivated by the desire to avoid withdrawal from xylazine, which several participants reported as well. However, in our qualitative analysis of posts on Reddit, we found a small number of participants that reported positive sentiments towards xylazine, indicating that there are some desirable effects of xylazine for some PWUDs, such as making the “high” experience last longer.

Despite these findings, most participants reported concerning adverse effects of xylazine use, such as prolonged sedation and “passing out” following use.^26^ As there are not currently any recommended reversal agents for xylazine, prolonged sedation is a clinical concern as patients who experience xylazine overdose may need prolonged respiratory or hemodynamic support.^1,4,25,26^ It is unclear if this prolonged sedation or synergistic respiratory depressant effect may be contributing to rising rates of opioid deaths and increasing prevalence of xylazine in overdose deaths.^25,27^ Some participants also reported an additional association of increased skin wounds with xylazine use. The biologic plausibility of xylazine use contributing to increased skin wounds has not been elucidated, however, this study adds to other studies that have reported this finding. ^1,13,14,28^

Second, this study corroborates the findings from other studies that xylazine use is concentrated in the northeastern United States.^14,27,28^ A majority of our participants reported that they were from the Northeastern United States; Pennsylvania was the most common state reported in that region. This is similar to the findings of Friedman et al. that xylazine use emerged in Philadelphia, Pennsylvania.^14^ Our study found that a majority of participants who used xylazine were younger (30-39 years old), non-Hispanic white, and male. Given the high rates of concurrent fentanyl use, it would be expected that xylazine use matches the demographics of people who use fentanyl. More recent data shows that fentanyl overdose death rates are more rapidly increasing among non-Hispanic African American males.^29^ This difference may reflect the demographics of people who use Reddit, who tend to be young, white, males.^17^ However continued research and surveillance is needed to assess the prevalence of xylazine in the drug supply in order to understand the burden of xylazine exposure among different racial, ethnic, age, and gender groups.^15,26^

Third, we found the PWUD reported worse withdrawal symptoms from other drugs after they had started using xylazine. Although we do not know the exact relationship between xylazine use and withdrawal from other drugs, this self-reported experience is concerning. Opioid Use Disorder has high morbidity and mortality, and ongoing withdrawal symptoms can lead to discontinuation of or further challenges to initiation of MOUDs.^30^ At this time the optimal management of xylazine withdrawal is unknown, although a case reports have described using clonidine or dexmedetomidine.^1,25,31^

Fourth this study demonstrates the potential opportunity to recruit participants from Reddit to participate in biomedical studies. Although there are a growing number of studies that use NLP to look at existing posts on Reddit to answer research questions, there are few studies that use Reddit for surveys or as a recruitment tool.^32–36^ Given the anonymity of Reddit and its large subscriber base, it could be a valuable source to survey questions about sensitive topics, perform toxicosurveillance, and study use patterns and adverse effects of emerging drugs with a novel method.^36^ As new psychoactive substances emerge in the drug market, Reddit could be a promising resource to study these substances with the goal of informing public health, harm reduction efforts, and clinical care.

Although this study did not directly answer the question of how harm from xylazine could be reduced, it highlights several issues facing PWUDs who are exposed to xylazine. More research is needed to understand the concomitant use of fentanyl and xylazine and the relationship between xylazine use and skin wounds.

### Limitations

There are several limitations to this study. As discussed above, the demographics of people who use Reddit may not be representative of the general population or the population of PWUDs, limiting the generalizability of findings in our study to all PWUDs.^17,35^ As telephone and internet access may be limited among PWUDs, many may not be able to utilize social media sites.^37^ Additionally, response bias could lead to people with polarized sentiments or who have had more reactions to xylazine to complete the survey, leading to an overestimate of people who had positive or negative reactions to xylazine. While this study’s findings that people who used xylazine tended to also use fentanyl, experience prolonged sedation, and had increased skin wounds is similar to other studies of people who use xylazine, more research is needed to understand the range of adverse effects of xylazine use.^1,13,25,27^ As the prevalence of xylazine in the drug supply moves across different geographic regions in the country, the demographics of its use concurrently with other drugs may change. Thus ongoing research into how xylazine is being used across time and geographics is needed to address the emerging needs of PWUDs who are exposed to xylazine.

## Conclusions

Among people on Reddit, xylazine appears to be an unwanted adulterant. PWUD may be experiencing more negative side effects such as prolonged sedation, increased skin wounds, and worsening withdrawal from other substances. This appeared to be more common problem in the Northeast and among people who self-report opioid use. Reddit shows promise as an avenue to disseminate surveys and possibly even recruit participants to studies that wish to explore sensitive topics.

## Data Availability

Reddit data is publicly available. The survey results are not available to the public.

