## Supplementary material for "Self-reported Xylazine Experiences: A Mixed Methods Study of Reddit Subscribers"

Supplement S1: Survey

Q1 What are your drugs of choice on a typical day(select all boxes that apply)?

- Fentanyl (1)
- Heroin (2)
- Oral Opioids (such as Oxycodone, Hydrocodone, Codeine (3)
- Marijuana (Weed, Pot, Hash) (4)
- Synthetic Marijuana (K2, Spice) (5)
- Hallucinogens (LSD, magic mushrooms, DMT, Peyote) (6)
- Cocaine (Crack, Blow, Coke) (7)
- Methamphetamine (Meth, Crystal Meth, Crank, Ice) (8)
- Amphetamine (Adderall, Bennies, Speed) (9)
- Ketamine (Special K) (10)
- MDMA (Ecstasy, Molly) (11)
- PCP (Angel Dust) (12)
- Benzodiazepines (Xanax, Vallium, Ativan, Benzos, Bars) (13)
- Inhalants (Nitrous Oxide, Poppers, Whippets) (14)
- Other (15) ________________________________________________

Q2 Have you ever used xylazine or xylazine containing drugs?

- Yes, intentionally (1)
- Yes, by accident (2)
- No (3)
- I am not sure (4)

Q3 How often do you use xylazine (Tranq) ?

- Daily (1)
- 4-6 times a week (2)
- 2-3 times a week (3)
- Once a week (4)
- 1-4 times a month (5)
- A few times a year (6)
- Never (7)

Q4 How do you use xylazine (Tranq) (select all that apply)?

- Mixed with other drugs, injection (1)
- Mixed with other drugs, intranasal (snort) (2)
- Mixed with other drugs, inhalational (smoke) (3)
- Mixed with other drugs, oral (4)
- On its own, injection (5)
- On its own, intranasal (snort) (6)
- On its own, inhalational (smoke) (7)
- On its own, oral (8)
- Never (9)

Q5 Do you intentionally seek to buy drugs (ex. Heroin, cocaine, etc.) that contain xylazine (Tranq)?

- Yes (1)
- No (2)

Q6 How does using xylazine (Tranq) effect your experience of getting high (select all that apply) ?

- A worse high experience (1)
- About the same/no change (2)
- A better high experience (3)
- Longer duration of high (4)
- Shorter duration of high (5)
- Sleepier (6)
- Nauseous/Vomiting (7)
- Increased focus (8)
- Decreased focus (9)
- Itchy (10)

Q7 Have you ever experienced withdrawal from xylazine (Tranq)?

- Yes (1)
- No (2)

Q8 If you have experienced withdrawal from xylazine (Tranq), what symptoms did you have (select all that apply)?

- I have never experienced withdrawal from xylazine (Tranq) (1)
- Body Aches (2)
- Cravings for zylazine (Tranq) (3)
- Nausea/vomiting (4)
- Diarrhea (5)
- Sweating (6)
- Anxiety (7)
- Depressed mood (8)
- Inability to sleep (9)
- Inability to focus (10)
- Sleep too much (11)
- Itchy (11)
- Fever (12)
- Cravings for other drugs (13)

Q9 How do you think withdrawing from another drug has changed since you started using xylazine (Tranq)

- A lot worse (1)
- A little worse (2)
- About the same (3)
- A little better (4)
- A lot better (5)

Q10 Are there side effects you have experienced from xylazine (Tranq) (select all that apply)?

- Increased overdoses (1)
- Increased Emergency Room (ER) visits (2)
- Increased wounds/abscesses (3)
- Increased need for naloxone (Narcan) (4)
- Increased passing out (5)

Q11 Are there any other side effects you have experienced from xylazine (Tranq) that you would like us, other researchers, or medical professionals to know about?

________________________________________________________________

Q12 Is there anything else about your experience with xylazine (Tranq) that you would like us, other researchers, or medical professionals to know about ?

________________________________________________________________

Q13 What is your age?

- Under 20 (1)
- 20-29 (2)
- 30-39 (3)
- 40-49 (4)
- 50-59 (5)
- Over 60 (6)

Q14 What gender do you identify with?

- Male (1)
- Female (2)
- Non-binary / third gender (3)
- Other (4) ________________________________________________
- Prefer not to say (5)

Q15 Are you of Hispanic, Latino, or Spanish origin such as Mexican, Puerto Rican, or Cuban?

- Yes (1)
- No (2)

Q16 Choose one or more races that you consider yourself to be:

- White (1)
- Black or African American (2)
- American Indian or Alaska Native (3)
- Asian (4)
- Native Hawaiian or Pacific Islander (5)
- Other (6) ________________________________________________

Q17 In which country do you currently reside?

▼ Afghanistan (1) ... Zimbabwe (1357)

Display This Question:

If List of Countries = United States of America

Q18 In which state do you currently reside?

▼ Alabama (1) ... I do not reside in the United States (53)

S2: Sample Reddit Post to recruit participants

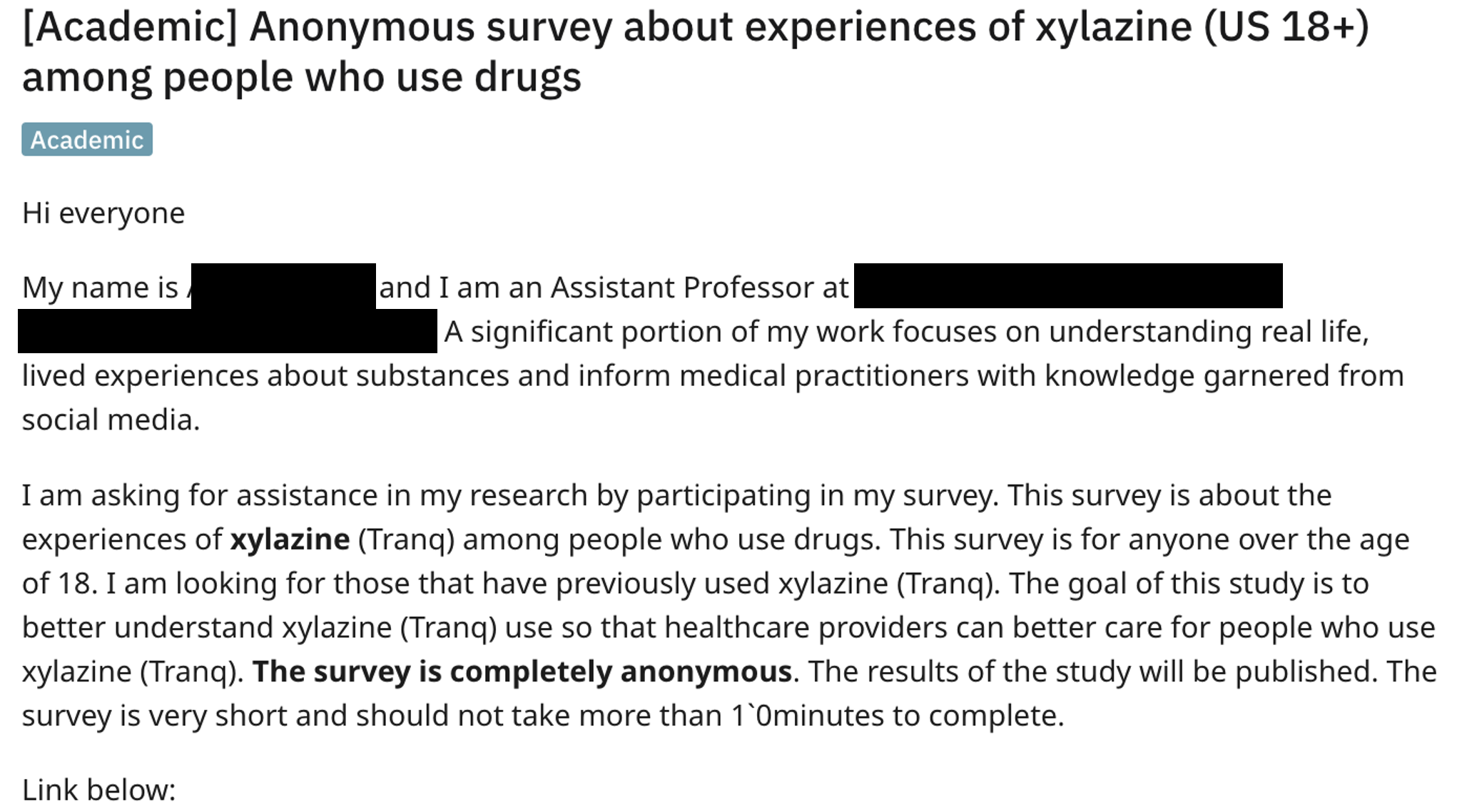
